# Lifetime Intimate Partner Violence (IPV) Against Mozambican Women: Individual and Contextual Level Factors Driving Its Prevalence

**DOI:** 10.1101/2024.10.12.24315379

**Authors:** Maud Z Muosieyiri, Nazeem Muhajarine

## Abstract

**Background:** Intimate partner violence (IPV) remains a significant public health issue in Mozambique. This study uses data from the 2022-2023 Mozambique Demographic and Health Survey (DHS) to examine the prevalence and sociodemographic determinants of Lifetime IPV among women.

**Methods:** A nationally representative sample of 4,813 women aged 15-49 was analyzed to assess the prevalence of Lifetime IPV. Logistic regression models were used to identify individual- and contextual-level factors associated with Lifetime IPV.

**Results:** Nearly 1 in 4 women (23.07%) reported experiencing physical abuse from a current or former partner in their lifetime. Marital status emerged as a key individual-level determinant, with married, cohabitating, and separated women being at significantly higher odds of experiencing IPV compared to women who had never been in a union. Educational attainment and current employment were also associated with increased odds of IPV. Similarly, women who justified physical abuse had higher odds of experiencing IPV. Additionally, husbands/partners’ alcohol consumption was one of the strongest predictors, nearly tripling the odds of Lifetime IPV. Finally, the effect modification between marital status and education showed that the intersection of these factors further shaped IPV risk. At the contextual level, provincial disparities were observed, with Cabo Delgado and Manica showing the highest IPV prevalence, while Inhambane and Gaza had the lowest.

**Conclusion:** This study provides updated data on the prevalence of Lifetime IPV in Mozambique and highlights key individual and contextual factors contributing to IPV. The findings underscore the need for targeted interventions addressing socio-cultural norms, improving educational opportunities, mitigating alcohol consumption, and implementing province-specific strategies to reduce IPV and enhance women’s safety across Mozambique.

## Background

Intimate Partner Violence (IPV) remains a major public health, social and moral issue affecting women worldwide. Addressing IPV is therefore key to achieving Sustainable Development Goals (SDGs) on gender equity (SDG 5), quality education (SDG 4), and health and wellbeing (SDG 3). According to the World Health Organization (WHO), nearly 27% of women aged 15 to 49 who have ever been in a relationship report having experienced physical and/or sexual violence by their intimate partner.(1) Physical violence perpetrated by a partner leads to multiple adverse health outcomes in women, for example, injuries, trauma, mental health effects, chronic conditions, and, among pregnant women, pregnancy complications like miscarriages and pre-term births.(1–3) Although IPV is prevalent across all societies and cultures, reportedly, it is highest within Latin America (31%), South-East Asia (33%), and Sub-Saharan Africa (SSA) (33%).(1,4)

In Sub-Saharan Africa, IPV prevalence differs by geographic location, likely due to variations in socio-cultural beliefs, gender norms, and economic inequality. For instance, a multi-country study in SSA showed that 50% of women in Ethiopia reported having experienced IPV in their lifetime, while only 17% of women in Namibia had ever experienced IPV.(5,6) Another pooled analysis of 26 African countries found that while 14% of pregnant women in South Africa having experienced physical violence during pregnancy, only 2.1% of those in Burkina Faso reported experiencing the same.(7) Ahinkorah et al., (2023) and colleagues determined that Gabon had the highest prevalence of IPV against women (45.3%), while Comoros had the lowest prevalence (4.9%) in their study of 84,486 women across 18 countries.(3) It is clear, given these reported data from across numerous countries, that understanding and addressing the specific socio-cultural, economic and gendered factors driving IPV at individual and contextual levels is essential for creating safer societies for women across Africa.(4,8)

African cultural traditions and gender norms widely reinforce men’s dominance as breadwinners and decision-makers in intimate partner relationships, making women more vulnerable to unjust treatment.(9,10) These norms intersect and interact with individual characteristics such as marital status, education, employment, attitudes towards abuse, and substance use; this shapes women’s susceptibility to, or protection from, IPV. The prevailing literature demonstrates that cohabitating/married women have a higher likelihood of experiencing IPV than their single counterparts. Cruz et al., (2014) revealed that cohabitating/married women had 1.53 times higher odds of experiencing physical violence compared to single women (OR = 2.53, 95% CI = 1.22, 4.74).(6) Ahinkorah and colleagues, in a notable finding, reported that cohabitating women are more likely to experience IPV than married women; possibly because women often may compromise more once they are married or feel that they have fewer options, resulting in less conflict with their husbands.(3)

Evidence also suggests that a woman’s higher level of education and employment, both general markers of women empowerment, can either reduce susceptibility to or trigger IPV, depending on the context.(11,12) On one hand, some studies show that higher education level and employment reduce the odds of IPV by increasing social networks and support, sources of information, leading to greater autonomy and better bargaining power in relationships.(3,4,8,13) For example, a study analyzing IPV among women across 16 Indian states determined that women with a higher than secondary education were 59% less likely to experience IPV compared to women with no formal education (OR = 0.41, 95% CI = 0.36, 0.46).(13) In contrast, in communities with rigid gender roles and acceptable views of and relaxed attitudes about physical violence, women’s higher education and employment increase their risk of experiencing IPV.(8) Cools and Kotsadam revealed that African women who achieved either a primary or a secondary education were significantly more likely to experience IPV, compared to those without formal education, the likelihood rising by 5.3 and 3.1 percentage points, respectively... Ahinkorah and colleagues (2018) showed that employed women had 33% higher odds of experiencing physical violence from their husbands/partners (OR = 1.33, 95% CI = 1.28, 1.37) compared to non-employed women.(4,7)

Researchers have hypothesized that women’s empowerment and IPV occur because empowered women challenge traditional, unfavourable (to women) gender roles, including questioning male authority.(8,14) Additionally, women’s attitudes towards IPV shape their vulnerability to experiencing it.(15) A population-based survey showed that IPV justification trends align with global IPV prevalence, with higher justification rates in South and Southeast Asia (47%) and Sub-Saharan Africa (38%) compared to Central and West Asia and Europe (29%).(15,16) Another study of African countries showed that pregnant women who justified IPV had a higher likelihood of experiencing physical violence than those who did not.(7) Scientists argue that in societies where women justify IPV, they are less likely to oppose it or to report it, thus increasing its occurrence.(17) Furthermore, alcohol use and abuse by women’s husbands/partners are consistently linked to higher rates of IPV.(18) Alcohol consumption can lower inhibitions, impair functioning, and heighten depressive symptoms, all of which may increase the likelihood of violence against women.(7)

At the contextual level, access to resource and wealth (as measured by wealth index), and rural/urban living have been reported to influence IPV trends. The Wealth Index is an aggregate score that measures the relative wealth of household’s wealth and may serve as a proxy for socioeconomic status.(3,19) Generally, women in the poorest wealth status are more likely to experience IPV than those in the richest category.(3,8) Stockl et al. showed that there is a significant decrease in the odds of experiencing IPV in richer households, compared to those in the middle and poorest tertile of the wealth measurement.(8) A strong supporting argument is that women within the richest wealth group are more resourced to fight for their rights and seek help against physical abuse compared to those of the poorest index.(7) Another argument is that financial stress is likely to be lesser reason for conflict in these well-endowed households.(3,8) There is mixed evidence concerning the influence of rural/urban living and IPV prevalence. Some studies show that rural living increases the odds of IPV due to rigid gender norms,(6) while others suggest that living in rural settings decreases IPV risk since women in these settings maybe more subservient, thus reducing any resistance to their husbands/partners dominance and aggression(7,17,20)

Mozambique, a southeastern African country, has historically had one of the continent’s highest IPV prevalence; the 2011 DHS report indicated that 33% of women had experienced physical violence since age 15.(17) Nevertheless, research suggests that this statistic is lower than the actual estimate due to underreporting, since IPV is often seen as a private issue, discouraging women from reporting violence and seeking support.(6,17,20) For instance, a 2011 study conducted in Zambezia, a central province in Mozambique, portrayed that 70% of participants admitted they never sought help or disclosed incidents of violence against them.(17,21) Despite historical reports, current data on IPV and its structural and sociodemographic drivers are largely unknown in Mozambique. This data gap makes it challenging to develop and implement effective policies and protocols to ensure the welfare and safety of women across the country. Nationally, the Mozambique Constitution establishes gender equality in all areas of society and prohibits all legislative, political, cultural, economic, and social discrimination. Many state bodies tasked with preventing and ending gender-based violence exists. However, lack of reliable and country-wide contemporary data hampers evidence-based action on this front. This present study, therefore, sought to address this evidence gap by utilizing the most recent Mozambique DHS data (2022-2023) to investigate the Lifetime IPV prevalence and its associated sociodemographic and structural factors among 15- to 49-year-old women in Mozambique.

## Methods

### Data Source

The present study analyzes information from a secondary data source, the 2022-2023 Mozambique DHS data.(19) The DHS, funded by the United States Agency for International Development (USAID), is a global survey that is conducted in over 85 low- and middle-income countries. We received approval from the DHS program to access de-identified datasets for the 2022-2023 Mozambique reports, which were provided on June 17, 2024. Thus, participant confidentiality was maintained throughout our analyses, and no information can be directly linked to any individual.

The 2022-23 Mozambique DHS was a nationwide, population-based cross-sectional survey that covered all 10 provinces (Niassa, Cabo Delgado, Nampula, Zambezia, Tete, Manica, Sofala, Inhambane, Gaza, and Maputo) as well as the capital city region of Maputo, which holds provincial status. Data collection followed a two-stage stratified sampling design. In the first stage, clusters (enumeration areas, EAs) were selected based on “IV Recenseamento Geral da População e Habitação 2017” (IV RGPH 2017)).(22) A total of 619 recorded areas were chosen using probability-proportional-to-size, determined by the number of households in each explicit stratum. In the second stage, 26 households were systematically selected equally from each area. This process resulted in the selection of 16,045 households for data collection. All women aged 15–49 years who were either residents or visitors in the household the night before the interviews were eligible to participate. In a subsample of half of the selected households, all men aged 15–54 years were also eligible for interviews.(19) For our analyses, we utilized data from a sub-group of women within the Individual Women Recode (IR) file who were randomly selected to complete the Domestic Violence module (N=4,813).

### Variables

#### Outcome Variable

We define the outcome variable, Lifetime Intimate Partner Violence (IPV), as any physical violence experienced by women from a current or former partner since the age of 15. This variable was derived from multiple questions according to the guidelines within the DHS-8 Guide to Statistics Manual.(23) A “Yes” response to any of the following questions met the criteria for Lifetime IPV: (a) Ever been pushed, shook or had something thrown by husband/partner; (b) Ever been slapped by husband/partner (c) Ever been punched with fist or hit by something harmful by husband/partner (d) Ever been kicked or dragged by husband/partner (e) Ever been strangled or burnt by husband/partner (f) Ever been attacked with knife/gun or other weapon by husband/partner (g) Ever CS physical violence by husband/partner (h) Ever had arm twisted or hair pulled by husband/partner (i) Previous husband: ever hit, slap, kick or physically hurt respondent. The outcome variable was dichotomized (“Yes”/ “No”) for all analyses.

#### Independent Variables

Seminal articles on IPV in Sub-Saharan Africa, like that of Ahinkorah et al., (2023) (7) as well as the adapted theoretical framework from Azevêdo et al (24) (**S1 Fig**), guided the selection of the independent variables. These variables were divided into Individual-level and Contextual-level factors. The Individual-level factors include (a) Maternal Age (b) Husband/Partner’s Age (c) Maternal Educational Level (d) Husband/Partner’s Educational Level (e) Woman’s Marital Status (f) Woman’s Current Employment Status (g) Woman’s Access to Media (h) Woman’s Justification for Beatings and (i) Husband/Partner’s Alcohol Intake. Contextual-level variables, but applied at individual level, comprised of (a) Wealth Index (b) Type of Place of Residence, and (c) Province of Residence. The supplemental material contains all the details of these variables, including their categorizations (**S2 Table**).

#### Data Analysis

Study population characteristics were summarized as counts and frequencies for all independent variables and presented in Table 1. Lifetime IPV (**Fig 1**) as well as IPV experienced within the last 12 months (**S3 Fig**) were also quantified as counts and frequencies and presented as figures. Additionally, binary logistic regression analysis was used to assess the association between Lifetime IPV and each independent variable, estimating the unadjusted odds ratios (OR) with their 95% confidence intervals (CI) (**S4 Table**). All variables with a bivariate association at p-value ≤ 0.20 were included in the multivariable logistic regression model in a stepwise process to achieve the most parsimonious final model. Selected (based on theoretical framework) variables were also tested for effect modifications. Finally, the Hosmer-Lemeshow test was used to determine the model’s goodness-of-fit. Because of smaller cell counts within the highest 2 categories of the variables: “Husband/Partner’s Age” and “Husband/Partner’s Educational Level”, these categories were merged to ensure sufficient sample in both the bivariate and multivariate analyses. All adjusted odds ratios, their 95% CIs, and corresponding p-values are reported in Table 2. The analyses were performed using SAS 9.4M8.

**Figure 1.**
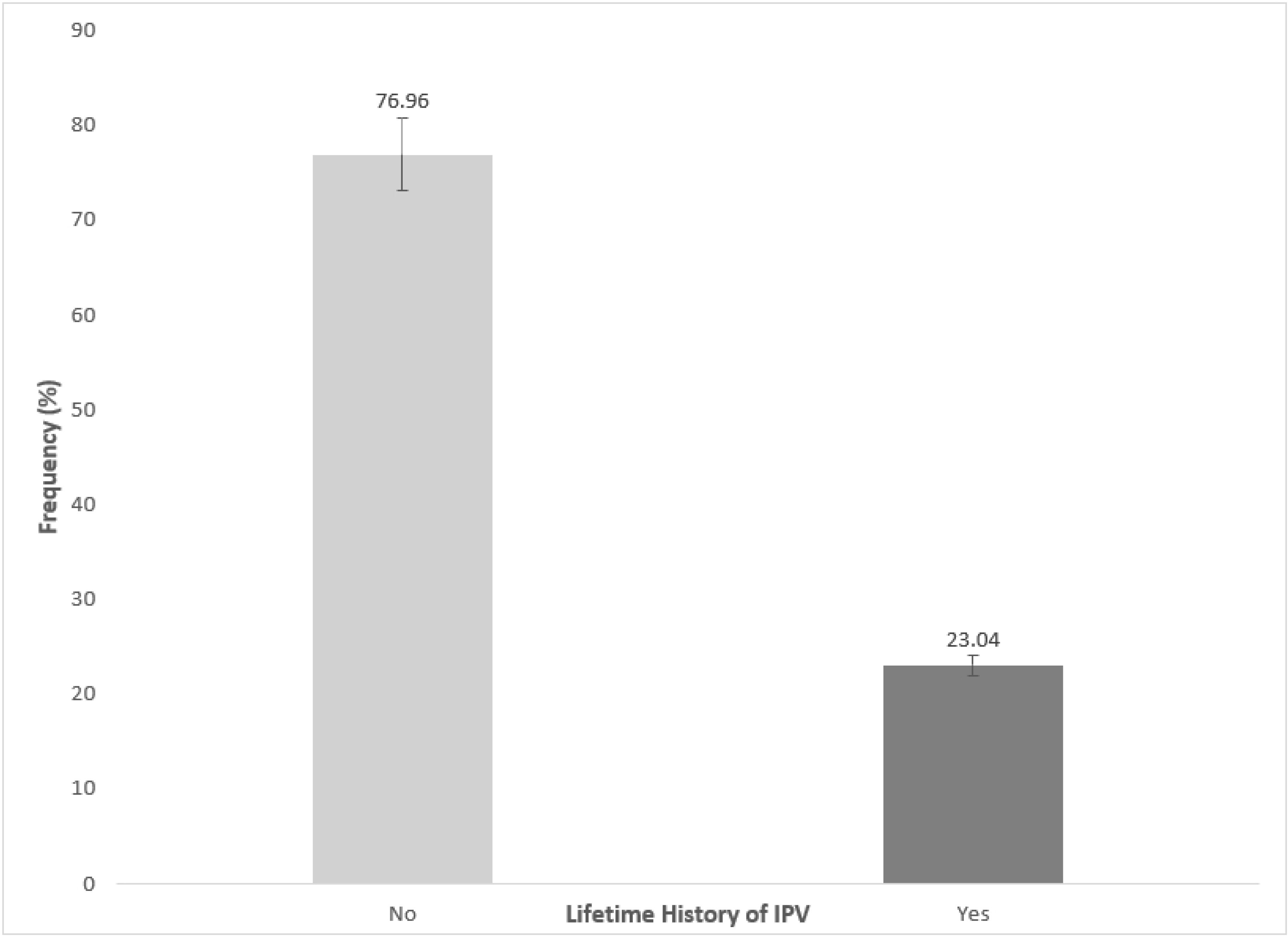
The prevalence of Lifetime Intimate Partner Violence (IPV) among women in Mozambique, based on the 2022-2023 DHS study, with a prevalence of 23.07% (N=4813)

**Table 1.** Background Information on Study Population (N=4,813). Demographic and Health Survey, 2022-2023, Mozambique.

| Variable | Frequency (N) | Frequency (%) |
| --- | --- | --- |
| **Woman's Age (N=4813)** |  |  |
| 15 – 24 | 1849 | 38.42 |
| 25 – 34 | 1501 | 31.19 |
| 35 – 44 | 1077 | 22.38 |
| ≥ 45 | 386 | 8.02 |
| **Husband/Partner's Age (N=3320)** |  |  |
| 15 – 24 | 442 | 13.31 |
| 25 – 34 | 1142 | 34.40 |
| 35 – 44 | 955 | 28.77 |
| ≥ 45 | 781 | 23.52 |
| **Woman's Marital Status (N=4813)** |  |  |
| Never in Union | 821 | 17.06 |
| Married | 1239 | 25.74 |
| Living with a Partner | 2081 | 43.24 |
| No longer living together/separated | 672 | 13.96 |
| **Woman's Educational Level (N=4813)** |  |  |
| No Formal Education | 1223 | 25.41 |
| Primary | 2062 | 42.84 |
| Secondary | 1371 | 28.49 |
| Higher | 157 | 3.26 |
| **Husband/Partner's Educational Level (N=3320)** |  |  |
| No Formal Education | 1025 | 30.87 |
| Primary | 1306 | 39.34 |
| Secondary | 856 | 25.78 |
| Higher | 133 | 4.01 |
| **Woman's Current Employment Status (N=4813)** |  |  |
| No | 3075 | 63.89 |
| Yes | 1738 | 36.11 |
| **Woman's Access to Media (N=4813)** |  |  |
| Less than Once a Week | 2872 | 59.67 |
| At Least Once a Week | 1941 | 40.33 |
| **Woman's Justification for Beating from Husband/Partner (N=4813)** |  |  |
| No justification | 3988 | 82.86 |
| Moderate Justification | 473 | 9.83 |
| Moderate-to-complete Justification | 352 | 7.31 |
| **Husband/Partner's Alcohol Consumption (N=4454)** |  |  |
| No | 2978 | 66.86 |
| Yes | 1476 | 33.14 |
| **Wealth Index (N=4813)** |  |  |
| Poorest | 776 | 16.12 |
| Poorer | 761 | 15.81 |
| Middle | 948 | 19.70 |
| Richer | 1089 | 22.63 |
| Richest | 1239 | 25.74 |

| **Type of Place of Residence (N=4813)** |  |  |
| --- | --- | --- |
| Rural | 2891 | 60.07 |
| Urban | 1922 | 39.93 |
| **Province of Residence (N=4813)** |  |  |
| Niassa | 439 | 9.12 |
| Cabo Delgado | 524 | 10.89 |
| Nampula | 552 | 11.47 |
| Zambézia | 376 | 7.81 |
| Tete | 469 | 9.74 |
| Manica | 405 | 8.41 |
| Sofala | 447 | 9.29 |
| Inhambane | 353 | 7.33 |
| Gaza | 413 | 8.58 |
| Maputo | 438 | 9.10 |
| Cidade de Maputo | 397 | 8.25 |

\*Some variables have missing data points, so each frequency is a proportion of the available data points for that question.
<sup>a</sup>Wealth Index: Composite score derived from participants' household assets using principal component analysis

**Table 2.**
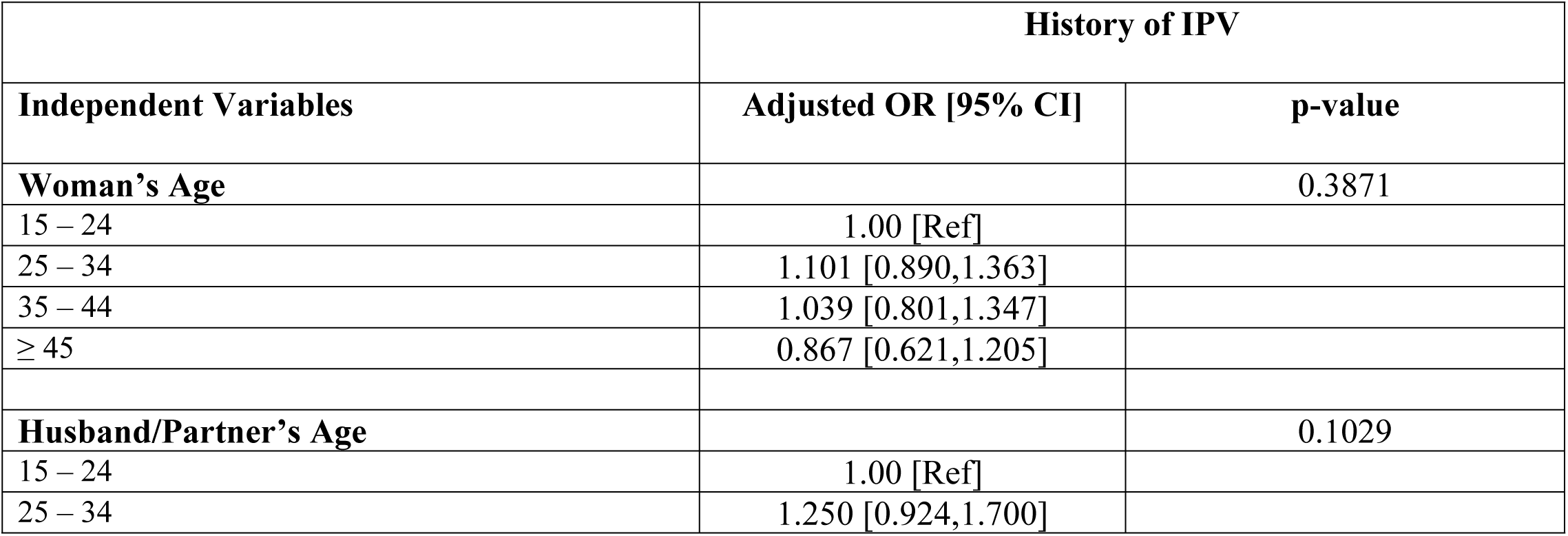
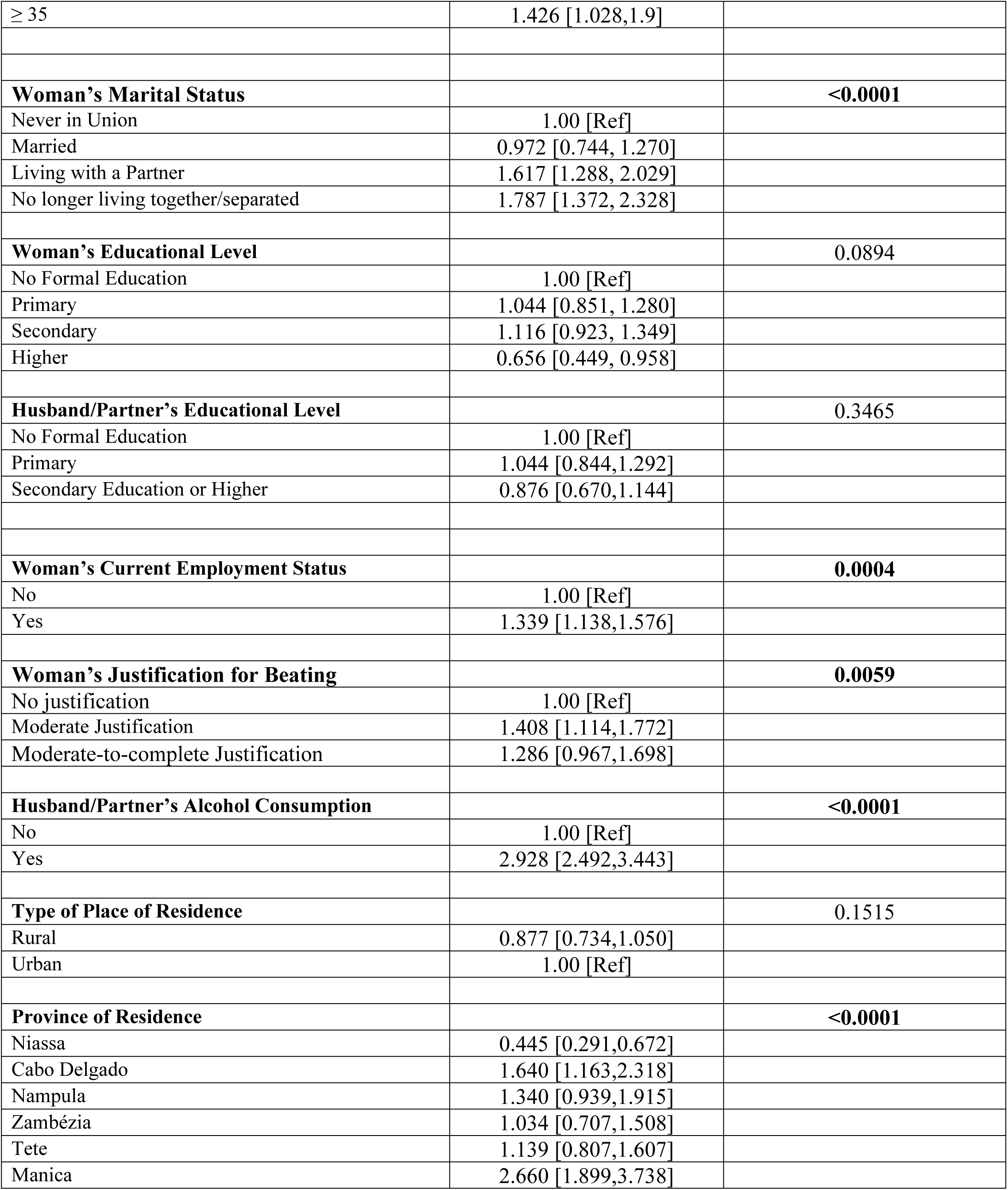
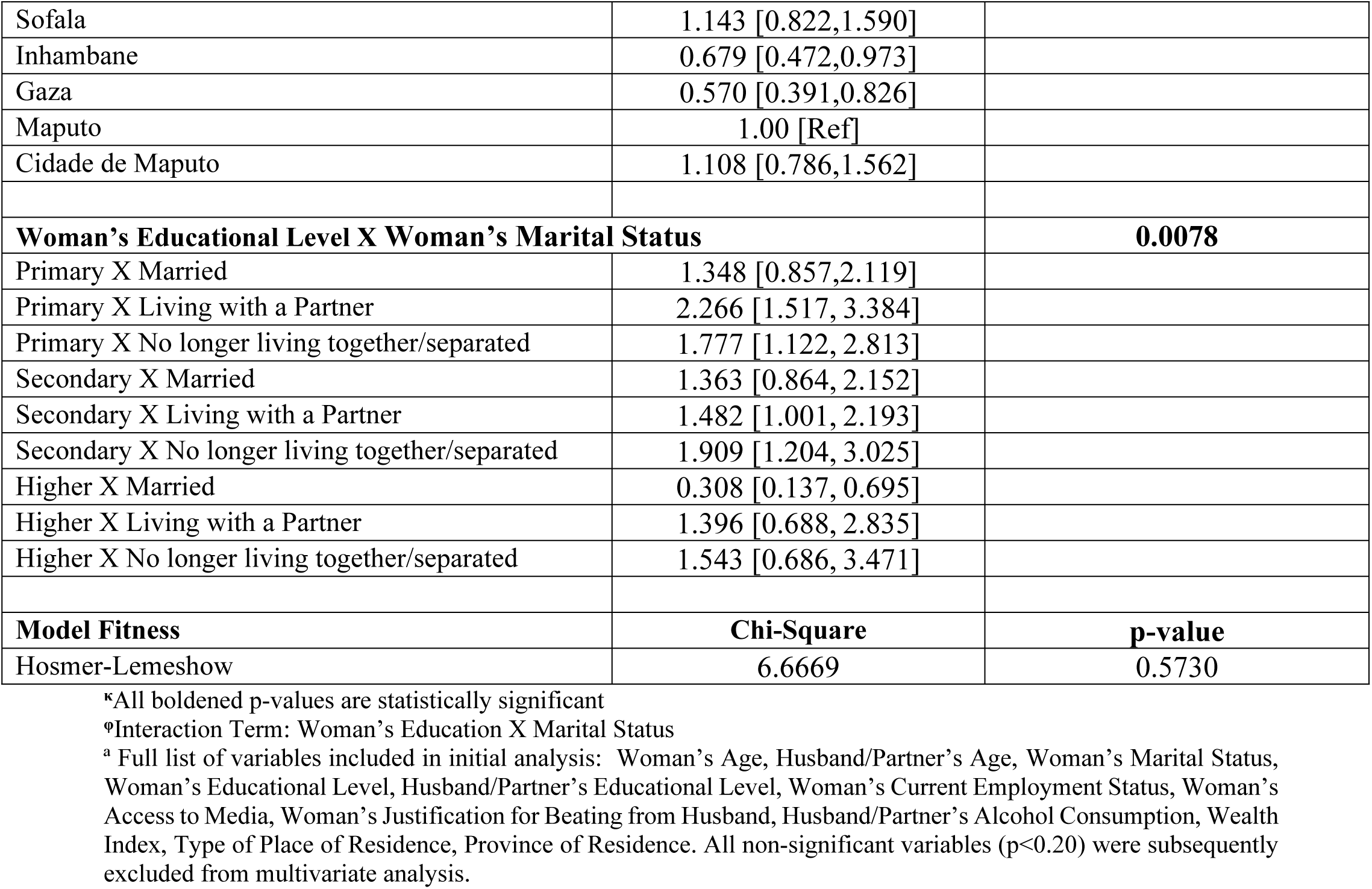
Results from the Multivariable Analysis, Full Model Showing Significant Factors at Individual- and Context-level and Lifetime Intimate Partner Violence. Demographic and Health Survey, 2022-2023, Mozambique.

## Results

### Sample Characteristics

Table 1 summarizes the background characteristics of the study population. The study population consisted of 4,813 women in Mozambique, with the largest proportion being between the ages of 15 and 24 (38.42%). In contrast, the majority of their husbands/partners were in the 25 to 34 age range (34.40%). Regarding marital status, most participants were either living with a partner (43.24%) or were married (25.74%). Educationally, the largest proportion of women (42.84%), and their husbands/partners (39.34%), had completed primary education, while only 3.26% and 4.01% had attained an education higher than secondary level, respectively. Concerning employment status, 63.89% of the women were unemployed at the time of the survey. Media access reported was limited, with more than half of the women reporting less than once-weekly access (59.67%). In terms of stating IPV was justified, the vast majority of women did not justify violence (82.86%). About one-third of the women stated that their husbands/partners (33.14%) consumed alcohol. Rural living was prevalent among the participants, with 60.07% residing in rural areas. The wealth distribution showed an even spread, with 16.12% in the poorest category and 25.74% in the richest category. Of the participants, 10.89% resided in Cabo Delgado, 11.47% in Nampula, and 9.12% in Niassa province.

### Prevalence of Lifetime IPV

Nearly 1 in 4 (23.07%) women reported that they had experienced physical abuse from a current or former partner since the age of 15 (**Fig 1**). This prevalence is similar to the IPV experienced within the preceding 12 months (21.34%) (**S3 Fig**). Figure 2 shows the breakdown of regional variation in Lifetime IPV across all provinces. Manica had the highest rate at 37.3%, followed by Cabo Delgado at 27.1% and Sofala at 28.4%. The lowest prevalence was found in Tete at 14.8% and Niassa at 9.6%.

**Figure 2.**
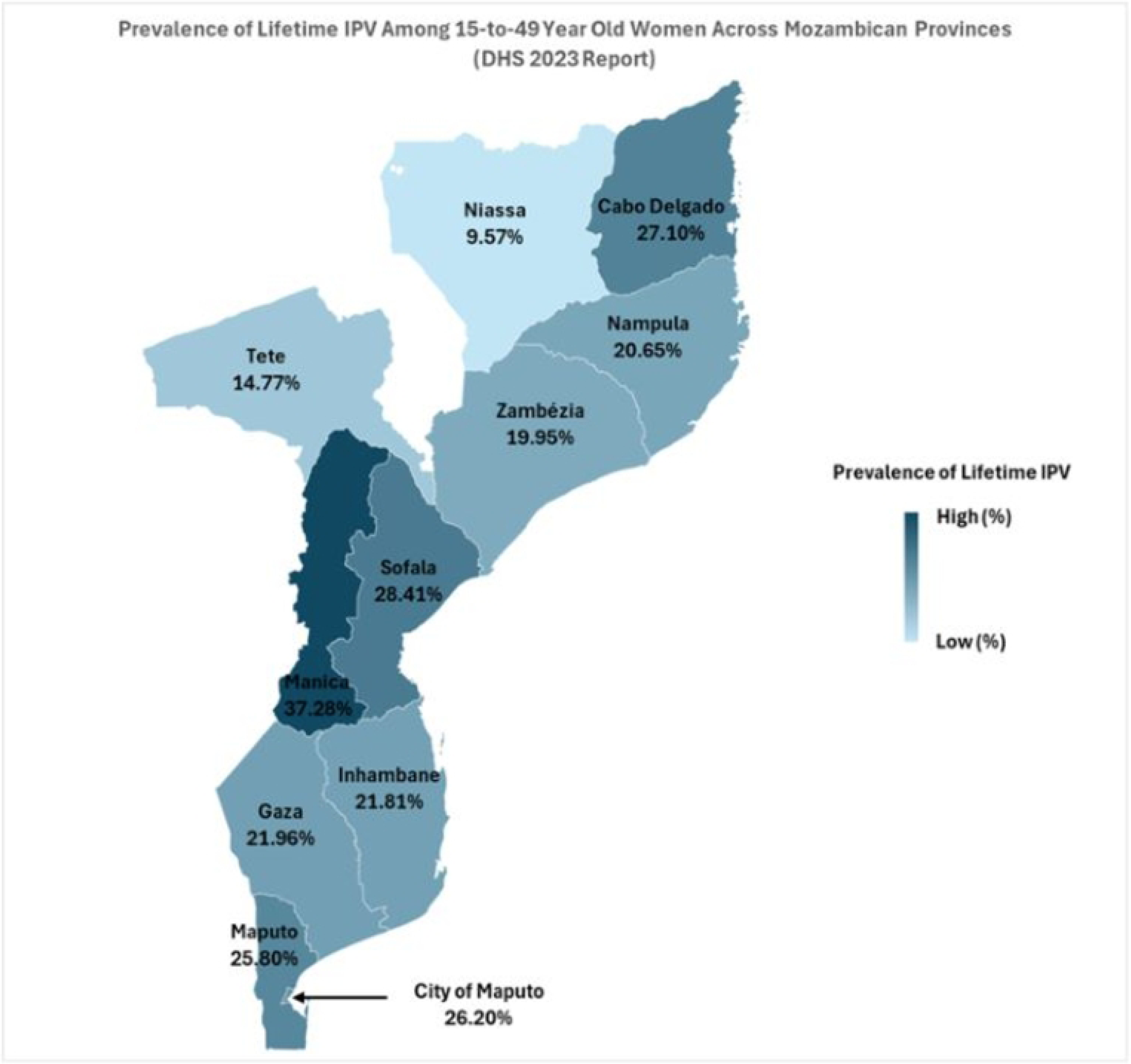
Provincial variation in the prevalence of Lifetime Intimate Partner Violence (IPV) among women in Mozambique, as reported in the 2022-2023 DHS study, highlighting the highest prevalence in Manica and the lowest in Niassa (N=4813)

### Individual-Level Factors Associated with the Prevalence of Lifetime IPV

Several individual-level factors were significantly associated with Lifetime IPV in this study (**Table 2**). Marital status was a strong determinant; women with a partner having a 61.7% higher likelihood of experiencing Lifetime IPV compared to those who had never been in a union (aOR 1.617, 95% CI 1.288, 2.029). Women who are no longer with a partner had 78.7% higher odds of Lifetime IPV (aOR: 1.787, 95% CI 1.372, 2.328). Education also played a role, women with higher than secondary education reporting decreased odds of Lifetime IPV (aOR: 0.656, 95% CI 0.449, 0.958). Employment status was another key factor; employed women had a 33.9% increased likelihood of experiencing Lifetime IPV compared to those not employed (aOR: 1.339, 95% CI 1.138, 1.576). Justification for beating was positively associated with Lifetime IPV; women expressing moderate degree of justification having 40.8% higher odds of Lifetime IPV (aOR: 1.408, 95% CI 1.114, 1.772) compared to those who did not justify violence. Husband/partner’s alcohol consumption was one of the strongest predictors. Husbands/partner’s alcohol drinking nearly tripled the odds of Lifetime IPV (aOR: 2.928, 95% CI 2.492, 3.443).

An effect modification involving women’s educational level and marital status was also observed. Women with primary education who were living with a partner (aOR: 2.266, 95% CI 1.517, 3.384) or separated (aOR: 1.777, 95% CI 1.122, 2.813) had significantly higher odds of Lifetime IPV compared to those with no formal education who had never been in a union. For women with secondary education, those living with a partner (aOR: 1.482, 95% CI 1.001, 2.193) or separated (aOR: 1.909, 95% CI 1.204, 3.025) also had increased odds of Lifetime IPV. Figure 3 further illustrates the interaction between women’s educational level and marital status and the associated probabilities of Lifetime IPV.

**Figure 3.**
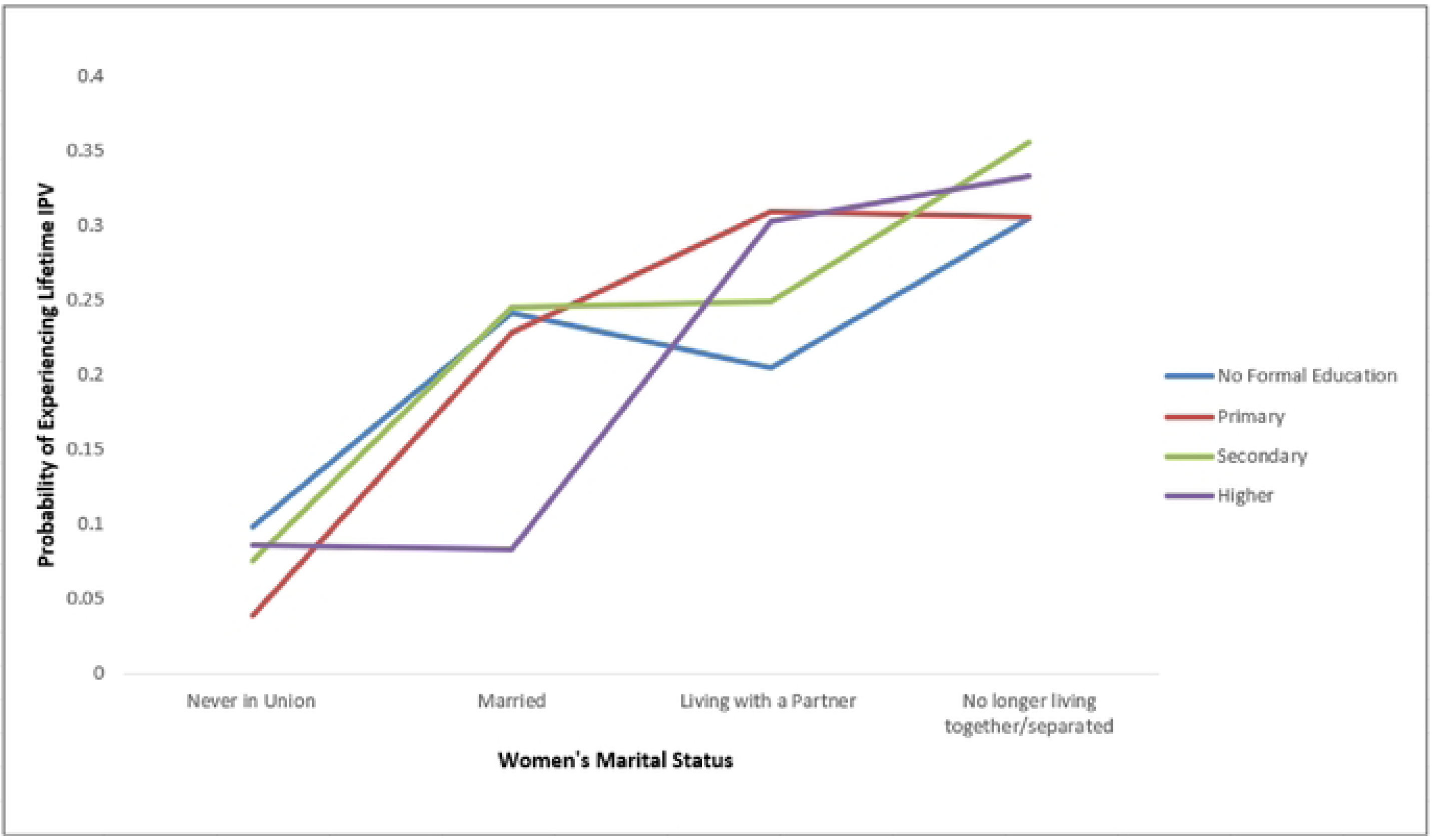
Interaction plot showing the predicted probabilities of experiencing Lifetime IPV across education levels and marital statuses. Higher education reduces the likelihood of Lifetime IPV, especially for married women, while those living with a partner or separated show consistently higher probabilities across all education levels.

For women with no formal education, those who have never been in a union show the lowest probability of IPV, while the highest probabilities are seen in those living with a partner or no longer living together. Among women with primary education, IPV probabilities are similar for those who are married or living with a partner, ranging from 0.22 to 0.30, while the gap between these groups and those never in union becomes more distinct. In secondary education, the probability of IPV increases slightly for married or partnered women, with separated women showing the highest IPV likelihood. For women with higher education, the probability of IPV decreases significantly for married individuals, but remains relatively high, around 0.30–0.33, for those living with a partner or separated. Overall, being married appears to reduce the probability of IPV as education increases, while the risks for those living with a partner or separated remain elevated across all education levels.

### Context-Level Factors Associated with the Prevalence of Lifetime IPV

The province of residence significantly impacted Lifetime IPV prevalence. Women from Cabo Delgado (aOR: 1.640, 95% CI 1.163, 2.318) and Manica (aOR: 2.660, 95% CI 1.899, 3.738) were at substantially higher odds of experiencing Lifetime IPV than those in Maputo. Conversely, women from Niassa (aOR: 0.445, 95% CI 0.291, 0.672), Inhambane (aOR: 0.679, [95% CI: 0.472, 0.973]), and Gaza (aOR: 0.570, 95% CI 0.391, 0.826) were less likely to experience IPV. No other context-level variables were significantly associated with Lifetime IPV. The Hosmer-Lemeshow test indicated a good fit for the logistic regression model (χ² = 6.67, p = 0.573).

## Discussion

Our study explored the prevalence and determinants of Lifetime IPV among women in Mozambique using data from the 2022-2023 Mozambique DHS. The study highlights key individual-level factors influencing IPV prevalence, including marital status, educational attainment, employment, justification of violence, and husband/partner’s alcohol consumption. We also found a significant effect modifier involving marital status and education level. On a contextual level, the study revealed regional disparities in IPV prevalence with women in Cabo Delgado and Manica reporting the highest IPV experiences.

Our study also revealed that marital status strongly influences the prevalence of Lifetime IPV. Women who were married, cohabitating, or separated were more likely to have experienced IPV in their lifetime than those who were not in a union. These findings align with the literature on this topic.(3,7,25) Studies show that IPV rates are higher among women with husbands/partners because men often use physical violence as a means to discipline as well as assert power and dominance over their wives/female partners.(25,26) Furthermore, women’s excessive reliance on their husbands/partners, in traditional Sub-Saharan African settings, further contributes to the high prevalence of IPV in certain cultures.(7,26)

Interestingly, our findings indicate that a woman’s education and employment are associated with a higher likelihood of experiencing IPV during her lifetime. This is notable since education and jobs are considered strong indicators of women’s autonomy and their financial and social power.(3,7,13) This somewhat counter-intuitive finding can be attributed to the deep-rooted rigid socio-cultural and gender norms found in many African countries.(3,8,15,17,20,25,27) As noted by Izugbara et al, educated and financially stable women may disrupt, even challenge, men’s traditional perceptions of their role as leaders, decision-makers, and primary providers in the household.(25) Thus, for men who strongly embrace conventional masculine norms, including male authority and dominance, this shift can heighten tensions and raise the risk of physical abuse.(25) Furthermore, some studies suggest that when husbands/partners are better educated or have higher-paying jobs, a woman’s education or occupation may not increase her risk of IPV, as power dynamics in this type of unions still favor men.(3,8,26) Our study however did not find any impact of husbands/partner’s education on IPV prevalence.

Consistent with existing literature, this study found that women’s justification of physical abuse significantly raised their odds of experiencing IPV.(7,15) One study estimates that women’s justification of physical abuse increases their likelihood of experiencing IPV by 30% while another study estimates a 57% higher odds of experiencing physical abuse.(7,25) A woman’s acceptance and justification of physical abuse are shaped by community attitudes that favor physical violence while dismissing female victimization. This context and environment not only increases the likelihood of being a victim but also pressures IPV victims to remain silent and accept the abuse.(4,15)

Another strong predictor of Lifetime IPV is a husband/partner’s alcohol consumption. We found that women whose husbands/partners consumed alcohol were more than twice as likely to experience IPV in their lifetime than those whose husbands/partners did not drink. Other studies have shown even stronger evidence that a husband/partner’s alcohol abuse increases the risk of experiencing abuse.(27–29) For instance, Olagbuji and colleagues found that having a partner who consumes alcohol raises the odds of experiencing IPV by 11 times.(29) Some sources suggest that alcohol consumption or addiction may lead men to neglect their families, increasing tensions in their intimate relationships that could result in physical abuse.(7) Others argue that alcohol consumption triggers immediate biological changes in men that lead to increased aggression and abuse toward their partners.(18)

Notably, the effect modification between marital status and education highlights how the intersection of these two individual-level factors can either heighten or mitigate the likelihood of experiencing IPV. This finding emphasizes the importance of recognizing how multiple dimensions of a woman’s identity and social position, intersect to shape her vulnerability to IPV in Mozambique. Higher educational attainment in women is typically linked to greater autonomy and resource access, which can reduce IPV risk.(30,31) However, in the contexts of strong traditional gender norms, educated women in intimate partner relationships might still greater vulnerability to experiencing IPV as their greater autonomy and independence challenge societal roles.(32) An intersectional approach provides a deeper understanding of these complexities and helps target effective interventions and policies.

On the contextual level, IPV prevalence varied by province, with Cabo Delgado and Manica having the highest statistics, and Inhambane and Gaza having the lowest. This disparity in IPV prevalence could be related to unequal wealth distribution across the country. According to a 2018 World Bank report, southern provinces like Inhambane, Gaza, Maputo, and Maputo City have smaller wealth distribution gaps between rural and urban areas and have a more even distribution of basic services, compared to others like Cabo Delgado, Manica, and Niassa.(33) Thus, unequal wealth and resource distribution in some provinces can worsen economic hardship, power imbalances, and psychological strain in relationships, increasing women’s vulnerability to IPV in their lifetime.

### Strengths and Limitations

The biggest strength of this study is that it utilized the most current data from a large, nationally representative dataset to provide a detailed analysis of Lifetime IPV prevalence, making our results generalizable to all women between 15 and 49 years in Mozambique. Additionally, our analysis effectively examined both individual- and contextual-level factors, therefore offering a holistic view of the phenomenon of IPV in the country. Furthermore, the study identified significant effect modification results between marital status and education, contributing to the understanding of how multiple dimensions of a woman’s identity affect IPV prevalence. Again, our study highlights important provincial variations in IPV prevalence, helping to identify high-risk areas of IPV that will guide tailored interventions accordingly.

Despite these strengths, some limitations exist. For example, the cross-sectional nature of the study prevents us from establishing a causal relationship between the independent variables and Lifetime IPV. Due to the stigma regarding IPV, some women may refuse to disclose their experience of abuse, leading to underreporting, and potentially skewing the final results. Furthermore, the use of self-report questionnaires could lead to non-differential misclassification and/or recall bias.

## Conclusion

Our study utilized the Mozambique DHS 2022-2023 data to examine the current prevalence of Lifetime IPV in the country and to identify the specific individual and contextual factors contributing to it. Our findings showed that almost 1 in 4 women experienced IPV in their lifetime. Marital status emerged as a key factor, with women who are currently married, cohabitating, or separated being at the highest odds of experiencing IPV in their lives. Educational attainment and current employment also played critical roles. Higher education levels and current employment correlated with increased IPV prevalence. Furthermore, the justification of violence significantly influenced IPV prevalence. Similarly, husbands/partners’ consumption of alcohol was strongly associated with Lifetime IPV prevalence. Finally, provincial disparities in Lifetime IPV were evident, with notably higher IPV estimates in Cabo Delgado and Manica.

Overall, our findings underscore the importance of targeted interventions that address socio-cultural norms, improve educational opportunities, mitigate alcohol consumption, and implement province-specific strategies to effectively combat Lifetime IPV and enhance women’s health and safety in Mozambique.

## Author Contributions

Conceptualization: MM and NM; Formal Analysis: MM and NM; Methodology: MM and NM; Supervision: NM; Funding and Provenance: NM; Writing – Original draft: MM. Writing – Review & Editing: NM.

## Data Availability

All relevant data are within the manuscript and its Supporting Information files.

## Acknowledgments

We thank Md Sabbir Ahmed, Nahin Shakurun, and Fernanda Andre for providing critical feedback during the conceptualization and manuscript writing processes of the project. We also appreciate the overall support and encouragement of all the Mozambique-Canada Maternal Health (MCMH) project colleagues.

## Supporting Information List

**S1 Figure.** The theoretical framework of physical violence against women during pregnancy. Four different levels contribute to the risk of physical violence during pregnancy. Level 1 includes contextual factors, Level 2 includes both a woman and her husband/partners socio-demographic indicators, Level 3 encompasses the dynamics within the relationship between a woman and her husband/partner, and Level 4 contains factors associated with the woman’s attitudes and intentions towards physical abuse. Adapted from intimate partner violence and unintended pregnancy framework by Azevêdo et al., (2013).

**S2 Table.** Details about Independent Variables and their Recode Status from Mozambique 20223-2023 Demographic and Health Survey Dataset

**S3 Figure.** The prevalence of Intimate Partner Violence (IPV) in the last 12 months among women in Mozambique, based on the 2022-2023 Demographic and Health Survey, with a prevalence of 21.34% (N=4813)

**S4 Table.** Results from Bivariate Analyses Between All Independent Variables and History of Intimate Partner Violence. Demographic and Health Survey, 2022-2023, Mozambique

